# High Throughput Deep Learning Detection of Mitral Regurgitation

**DOI:** 10.1101/2024.02.08.24302547

**Authors:** Amey Vrudhula, Grant Duffy, Milos Vukadinovic, David Liang, Susan Cheng, David Ouyang

## Abstract

**Background:** Diagnosis of mitral regurgitation (MR) requires careful evaluation of echocardiography with Doppler imaging. This study presents the development and validation of a fully automated deep learning pipeline for identifying apical-4-chamber view videos with color Doppler and detection of clinically significant (moderate or severe) mitral regurgitation from transthoracic echocardiography studies.

**Methods:** A total of 58,614 studies (2,587,538 videos) from Cedars-Sinai Medical Center (CSMC) were used to develop and test an automated pipeline to identify apical-4-chamber view videos with color Doppler across the mitral valve and then assess mitral valve regurgitation severity. The model was tested on an internal test set of 1,800 studies (80,833 videos) from CSMC and externally evaluated in a geographically distinct cohort of 915 studies (46,890 videos) from Stanford Healthcare (SHC).

**Results:** In the held-out CSMC test set, the view classifier demonstrated an AUC of 0.998 (0.998 - 0.999) and correctly identified 3,452 of 3,539 MR color Doppler videos (sensitivity of 0.975 (0.968-0.982) and specificity of 0.999 (0.999-0.999) compared with manually curated videos). In the external test cohort from SHC, the view classifier correctly identified 1,051 of 1,055 MR color Doppler videos (sensitivity of 0.996 (0.990 – 1.000) and specificity of 0.999 (0.999 – 0.999) compared with manually curated videos). For evaluating clinically significant MR, in the CSMC test cohort, moderate-or-severe MR was detected with AUC of 0.916 (0.899 - 0.932) and severe MR was detected with an AUC of 0.934 (0.913 - 0.953). In the SHC test cohort, the model detected moderate-or-severe MR with an AUC of 0.951 (0.924 - 0.973) and severe MR with an AUC of 0.969 (0.946 - 0.987).

**Conclusions:** In this study, we developed and validated an automated pipeline for identifying clinically significant MR from transthoracic echocardiography studies. Such an approach has potential for automated screening of MR and precision evaluation for surveillance.

## Introduction

Mitral regurgitation (MR) is one of the most common forms of valve disease, affecting more than 4 million Americans.^1,2,3^ Often progressing insidiously and frequently underrecognized^3^, both primary MR as well as secondary MR can be initially asymptomatic but lead to worsening heart failure and mortality.^1,4–6,7,8^ There has been an increased focus on early MR diagnosis given advances in surgical and transcatheter treatment options^1,9,10^. Echocardiography with color Doppler is the most common method of initial evaluation of MR, with a holistic assessment combining left atrial size, effective regurgitant orifice area, regurgitant fraction, regurgitant volume, as well as other key clinical factors to accurately assess disease severity.^11,12^ Despite ultrasound technology becoming more widely available, accurate assessment of MR still requires experienced expert image acquisition and evaluation.

Recent advances in machine learning offer opportunities to automate time-consuming steps in the interpretation of medical imaging. Artificial intelligence (AI) has the ability to precisely phenotype subtle cardiac physiology as well as identify imaging features of disease severity not recognized by clinicians.^13–16^ Deep learning has been applied to echocardiography to improve the precision of common measurements, such as left ventricular ejection fraction^13^ and wall thickness^15,17^, as well as streamlining assessment of aortic stenosis^18^, hypertrophic cardiomyopathy (HCM)^15^, and cardiac amyloidosis (CA).^15,19^ With increased ultrasound availability, AI guidance has been developed for both image acquisition and interpretation^13,20^. With the increasing prevalence of MR in an aging population with co-morbid heart failure, AI could aid in MR screening and surveillance.^21–25^

In this study, we developed and evaluated a deep learning pipeline’s performance in automating identification of MR from standard transthoracic echocardiogram studies. We hypothesized that a deep learning approach can identify color Doppler apical-4-chamber videos and assess MR severity with high-throughput automation, and this automated pipeline was evaluated in two geographically distinct cohorts (**Figure 1**). Combined with other echocardiography AI algorithms, such an approach can be used for serial surveillance and screening of mitral regurgitation.

**Figure 1:**
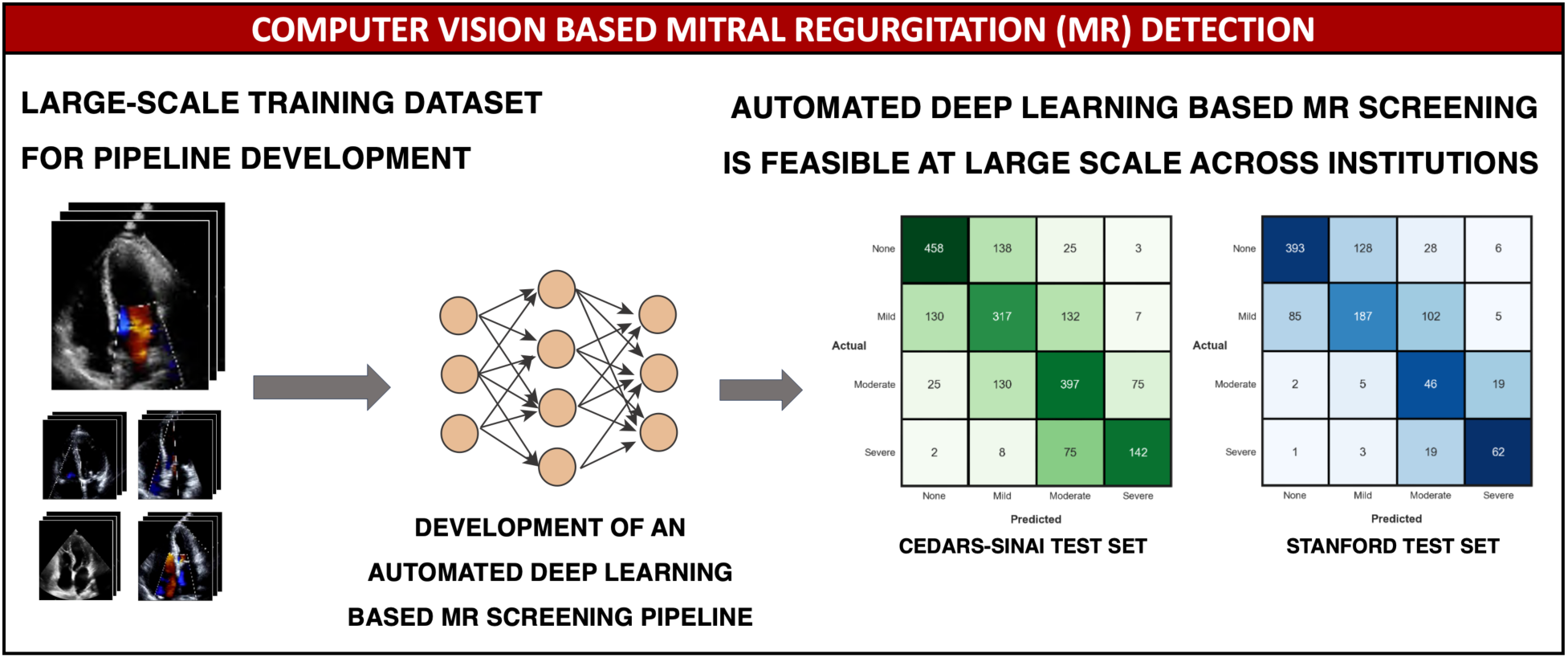
Computer Vision Based Mitral Regurgitation (MR) Detection: An automated deep learning pipeline was trained to detect and stratify mitral regurgitation severity using large-scale data consisting of apical-4-Chamber (A4C) echocardiogram videos with Color Doppler across the mitral valve (CSMC). The automated pipeline showed strong and consistent performance in test sets at CSMC and SHC. These results show that deep learning can accurately detect clinically significant MR using single-view TTE videos with Doppler information. Deep learning-based MR detection tools could serve as a part of point-of-care ultrasound screening as part of clinic visits or in resource limited settings where imaging may be obtained by individuals with minimal training.

## Methods

### Study Population and Data Source

#### Cedars-Sinai Medical Center (CSMC) Cohort

A total of 58,614 transthoracic echocardiogram studies from 38,461 patients receiving care at Cedars-Sinai Medical Center (CSMC) between October 11, 2011 and June 04, 2022 were used to train and evaluate the deep learning models. A total of 2,587,538 videos (an average of 44 videos per study after excluding still images) were initially sourced from Digital Imaging and Communications in Medicine (DICOM) files and underwent de-identification, view classification, and pre-processing into AVI videos. 354,117 videos were classified as apical-4-chamber videos using an automated view classifier, and then manually curated to identify 34,714 videos with color Doppler across the mitral valve.^26^

Following view selection, the CSMC cohort included 34,714 videos across 30,453 unique echocardiogram studies from 22,661 patients, and a subset enriched for moderate and severe MR were used for training. A total of 20,604 videos from 18,133 studies in the dataset were split on a patient level into train (80%), validation (10%), and test (10%) cohorts to train a deep neural network for MR severity classification (**Figure 2**). MR severity for each study was determined based on the clinical echocardiogram report determined in a high volume echocardiography lab in accordance with ASE guidelines.^27^ When MR was characterized as an intermediate category (“trace to mild” or “mild to moderate” or “moderate to severe”), videos were placed in the more severe categories. Both primary and secondary MR were included. Studies with concomitant mitral stenosis, other prosthetic valves, and heart failure were also included in both training and validation datasets.

**Figure 2:**
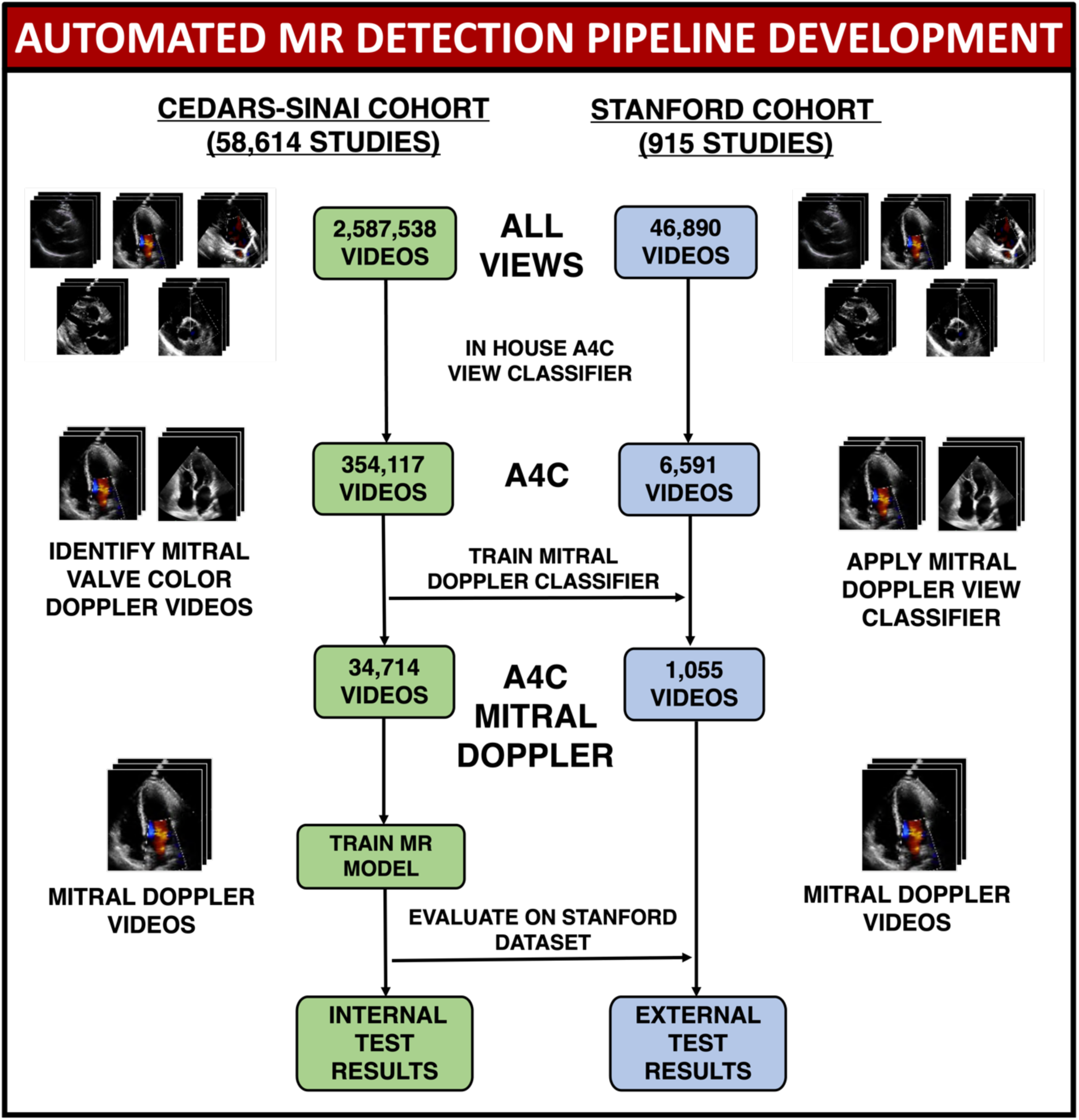
CSMC and Stanford Dataset Isolation. 34,714 Color Doppler A4C videos were isolated from a larger set of videos from CSMC. A view classifier was trained and used to isolate A4C Mitral Doppler videos from 915 studies containing 1,055 suitable videos from Stanford Healthcare. The MR classification model was then benchmarked on an internal test set from CSMC and an external test set from Stanford Healthcare.

#### Stanford Healthcare (SHC) Cohort

The pipeline was evaluated on 915 studies (containing a total of 46,890 videos) from SHC’s high-volume academic echocardiography lab. The automated view classification pipeline was compared with manual curation of videos within those studies to evaluate specificity. All videos identified by the view classifier were used for downstream MR severity model validation. Model output was compared with MR severity determined by expert cardiologists from the clinical reports. This study was approved by the Institutional Review Boards at Cedars-Sinai Medical Center and Stanford Healthcare. The need for informed consent was waived as the study involved secondary analysis of existing data.

### AI Model Training

Deep learning models were trained using the PyTorch Lightning deep learning framework. When patients had multiple echocardiogram videos and studies, each video was considered an independent example during training, with care not to have patient overlap across training, validation, and test cohorts. Video-based convolutional neural networks (R2+1D) were used for view classification and MR severity assessment.^28^ This model architecture was previously used for other echocardiography tasks and shown to be effective.^17^ The models were initialized with random weights and trained using a binary cross entropy loss function for up to 100 epochs, using an ADAM optimizer, an initial learning rate of 1e-2, and a batch size of 24 on two NVIDIA RTX 3090 GPUs. Early stopping was performed based on the validation loss.

The view classifier was trained using the 34,714 manually curated videos with color Doppler across the mitral valve as cases and 49,263 other apical-4-chamber videos as controls. Controls were a combination of videos that did not have color Doppler or had color Doppler window not focused on the mitral valve (videos focused on the tricuspid valve, intra-atrial septum, or ventricular septum). The MR severity model was trained on 6,206 videos without MR, 6,128 videos with mild MR, 6,174 videos with moderate MR and 2,042 videos with severe MR. This process is summarized in **Figure 2**.

### Statistical Analysis

Model performance was evaluated using area under the receiver operating characteristic curve (AUROC) and confusion matrices. F1-score, recall (sensitivity), positive predictive value (PPV), and negative predictive value (NPV) for both greater than moderate MR and severe MR. During external validation, the view classifier and MR classifier were evaluated serially as an automated pipeline. Statistical analysis was performed in Python (version 3.8.0) and R (version 4.2.2). Confidence intervals were computed via bootstrapping with 10,000 samples. Reporting of study results are consistent with guidelines put forth by CONSORT-AI.^29,30^

### Model Explainability

The key imaging features identified by the MR severity model were evaluated using saliency mapping generated using the Integrated Gradients method.^31^ This method generated a heatmap for every frame of the video, summarized as a final 2-dimensional heatmap generated by using the maximum value along the temporal axis for each pixel location in the video. Pixels brighter in intensity and closer to yellow were more salient to model predictions, while those darker in color were less important to the model’s final prediction. When assessing videos with no MR, heatmaps were obtained by taking the maximum of saliency maps for the moderate and severe class output neurons for each pixel location (**Figure 4**).

## Results

### Study Population

A total of 58,614 studies from 38,461 patients were used to train the deep learning pipeline. From 2,587,538 initial videos, a total of 354,117 videos were identified as apical-4-chamber and subsequently manually curated to identify videos that had color Doppler across the mitral valve. The manually curated color Doppler videos were used to train a view-classification model and linked with clinician reports to train the MR severity model. Patient characteristics are presented in **Tables 1 & 2** and are representative of the general CSMC patient population that received echocardiograms. The data was split on patient level for training and validation and had similar patient age, ejection fraction, left atrial volume index and proportions of male sex, coronary artery disease, and atrial fibrillation.

**Table 1 -.** Clinical and demographic characteristics represented in the training, validation, and internal test data sets for the 83,977 apical-4-chamber videos used to train, validate, and test the mitral doppler A4C view classifier. Values outside and inside parentheses represent number and percent, respectively, for categorical variables and mean and standard deviation for continuous variables.

|  | Train | Validation | Test |
| --- | --- | --- | --- |
| **Videos, n (%)** | 67,179 (80.0) | 8,344 (9.9) | 8,454 (10.1) |
| **Patients, n (%)** | 30,761 (80.0) | 3,848 (10.0) | 3,851 (10.0) |
| **Studies, n (%)** | 46,882 (80.0) | 5,790 (9.9) | 5,942 (10.1) |
| **Male, n (%)** | 38,367 (57.1) | 4,907 (58.8) | 4,854 (57.4) |
| **Hypertension, n (%)** | 40,982 (61.0) | 5,070 (60.8) | 5,065 (59.9) |
| **Coronary Artery Disease, n (%)** | 29,661 (44.2) | 3,670 (44.0) | 3,758 (44.5) |
| **Atrial Fibrillation, n (%)** | 19,909 (29.6) | 2,289 (27.4) | 2,498 (29.5) |
| **Ejection Fraction, mean (SD), %** | 56.56 (16.2) | 57.11 (15.5) | 55.85 (16.4) |
| **Left Atrial Volume Index (LAVI), mean (SD), mL/m<sup>2</sup>** | 34.71 (15.4) | 34.49 (15.4) | 34.05 (15.1) |
| **Race, n (%)** |  |  |  |
| White | 47,110 (70.1) | 5,927 (71.0) | 5,709 (67.5) |
| Black | 8,282 (12.3) | 972 (11.6) | 1,228 (14.5) |
| Asian | 5,232 (7.8) | 579 (6.9) | 640 (7.6) |
| Other | 6,555 (9.8) | 866 (10.4) | 877 (10.4) |
| **MR Severity, n (%)** |  |  |  |
| No MR | 24,840 (37.0) | 3,107 (37.2) | 3,291 (38.9) |
| Mild | 25,235 (37.6) | 3,143 (37.7) | 3,104 (36.7) |
| Moderate | 12,477 (18.6) | 1,519 (18.2) | 1,518 (18.0) |
| Severe | 4,627 (6.9) | 575 (6.9) | 541 (6.4) |

**Table 2 -.** Clinical and demographic characteristics of the 20,604 apical-4-chamber videos used to train, validate, and test the MR model. Values outside and inside parentheses represent number and percent, respectively, for categorical variables and mean and standard deviation for continuous variables.

|  | Train | Validation | Test |
| --- | --- | --- | --- |
| **Videos, n (%)** | 16,533 (80.2) | 2,007 (9.7) | 2,064 (10.0) |
| **Patients, n (%)** | 11,623 (80.0) | 1,450 (10.0) | 1,450 (10.0) |
| **Studies, n (%)** | 14,565 (80.3) | 1,768 (9.8) | 1,800 (9.9) |
| **Male, n (%)** | 9,628 (58.2) | 1,143 (57.0) | 1,152 (55.8) |
| **Hypertension, n (%)** | 10,087 (61.0) | 1,179 (58.7) | 1,296 (62.8) |
| **Coronary Artery Disease, n (%)** | 7,482 (45.3) | 888 (44.2) | 963 (46.7) |
| **Atrial Fibrillation, n (%)** | 5,414 (32.7) | 599 (29.8) | 642 (31.1) |
| **Ejection Fraction, mean (SD), %** | 54.53 (17.6) | 55.26 (17.5) | 54.0 (17.9) |
| **Left Atrial Volume Index (LAVI), mean (SD), mL/m<sup>2</sup>** | 36.85 (15.8) | 36.35 (15.8) | 37.44 (16.3) |
| **Race, n (%)** |  |  |  |
| White | 11,567 (70.0) | 1,378 (68.7) | 1,400 (67.8) |
| Black | 1,983 (12.0) | 277 (13.8) | 283 (13.7) |
| Asian | 1,428 (8.6) | 147 (7.3) | 192 (9.3) |
| Other | 1,555 (9.4) | 205 (10.2) | 189 (9.2) |
| **MR Severity, n (%)** |  |  |  |
| No MR | 4,949 (29.9) | 633 (31.5) | 624 (30.2) |
| Mild | 4,990 (30.2) | 606 (30.2) | 586 (28.4) |
| Moderate | 4,952 (30.0) | 595 (29.6) | 627 (30.4) |
| Severe | 1,642 (9.9) | 173 (8.6) | 227 (11.0) |

### View Classifier Performance Across Two Institutions

On a test set of 3,109 studies (132,767 videos) from CSMC not seen during model training, the view classifier identified 3,452 videos (97.5% of manually identified cases). This corresponds to an of AUC of 0.998 (0.998 – 0.999), and at the Youden Index, with a sensitivity of 0.975 (0.968 - 0.982) and specificity of 0.999 (0.999-0.999). To evaluate generalization of the view classification model at a geographically distinct site, we evaluated its performance on 915 studies from SHC. The view classifier isolated 1,091 videos from a total of 46,890 videos, while manual review identified 1,055 videos with color Doppler across the mitral valve. The view classifier correctly identified 1,051 (99.6%) of manually curated videos, with 4 videos not found by the AI pipeline and 40 false positives. This corresponds to a sensitivity of 0.996 (0.990 – 1.000) and specificity of 0.999 (0.999 – 0.999).

### Mitral Regurgitation Severity Performance Across Two Institutions

The MR severity model showed strong performance in distinguishing MR severity and identifying clinically significant mitral regurgitation (**Figure 3**). In the internal CSMC test set not used during model training, the model demonstrated an AUC of 0.916 (0.899 - 0.932) in detecting ≥ moderate MR and an AUC of 0.934 (0.913 - 0.953) for severe MR. The AI model had an NPV of 0.954 (0.940 - 0.967) for severe MR and an NPV of 0.863 (0.835 - 0.890) for ≥ moderate MR. Further information on MR model performance is presented in **Table 3**. The MR severity model performance was similar across institutions. In the SHC cohort, the model identified severe MR with an AUC of 0.969 (0.946 - 0.987) and ≥ moderate MR with an AUC of 0.951 (0.924 - 0.973). In this cohort, the model had an NPV of 0.977 (0.962 – 0.990) for severe MR and an NPV of 0.986 (0.974 – 0.995) for ≥ moderate MR.

**Figure 3:**
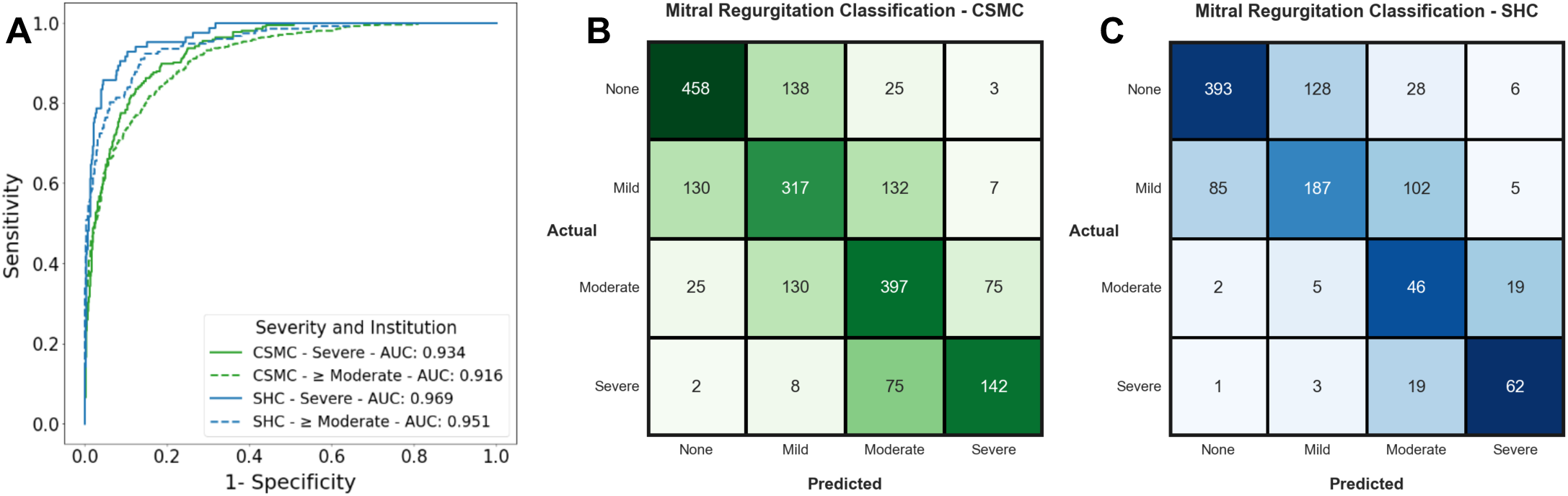
Model Performance Across Severity and Institution -. **A.** Receiver operating characteristic (ROC) curves for detection of Severe or ≥ Moderate MR at CSMC and Stanford. “≥ Moderate” included moderate, moderate to severe, and severe MR. **3B and 3C:** MR Classification on test set videos from CSMC and Stanford, respectively. Confusion matrix colormap values were scaled based on the proportion of actual disease cases in each class that were predicted in each possible disease category. This was done to allow for relative comparison of model performance across disease classes (None, Mild, Moderate, and Severe) given class imbalance.

**Table 3 -.** Model performance across institutions. - AUC, PPV, NPV, Recall and F1-score for MR on an internal test set from CSMC and an external validation set at Stanford. 95% confidence intervals were obtained by bootstrapping 10,000 samples. “≥ moderate” includes moderate and severe MR.

| <u>Site</u> | <u>Class</u> | <u>AUC</u> | <u>PPV</u> | <u>NPV</u> | <u>Recall</u> | <u>F1-Score</u> |
| --- | --- | --- | --- | --- | --- | --- |
| CSMC | ≥ Moderate MR | 0.916<br>(0.899 - 0.932) | 0.805<br>(0.766 - 0.842) | 0.863<br>(0.835 - 0.890) | 0.807<br>(0.769 - 0.843) | 0.806<br>(0.776 - 0.834) |
|  | Severe MR | 0.934<br>(0.913 - 0.953) | 0.626<br>(0.533 - 0.713) | 0.954<br>(0.940 - 0.967) | 0.626<br>(0.535 - 0.713) | 0.626<br>(0.546 - 0.695) |
| Stanford | ≥ Moderate MR | 0.951<br>(0.924 - 0.973) | 0.509<br>(0.427– 0.591) | 0.986<br>(0.974 – 0.995) | 0.930<br>(0.867 – 0.983) | 0.658<br>(0.581 – 0.727) |
|  | Severe MR | 0.969<br>(0.946 - 0.987) | 0.674<br>(0.537 – 0.809) | 0.977<br>(0.962 – 0.990) | 0.729<br>(0.580 – 0.860) | 0.701<br>(0.581 – 0.800) |

### Model Interpretation

Notably, saliency maps for our model demonstrate that the model focuses on the clinically relevant imaging features of mitral regurgitation. Saliency maps from Integrated Gradients were used to identify regions of interest in each video contributing the most to detection of MR severity (**Figure 4**).^31^ These interpretability techniques demonstrated localization of the activation signal in the color Doppler window and primarily highlighting the regurgitant jet, indicating that the model used appropriate, physiologic features of the mitral regurgitation to make predictions. Frame-by-frame saliency visualizations are shown in **Supplemental Videos S1-S4**.

**Figure 4:**
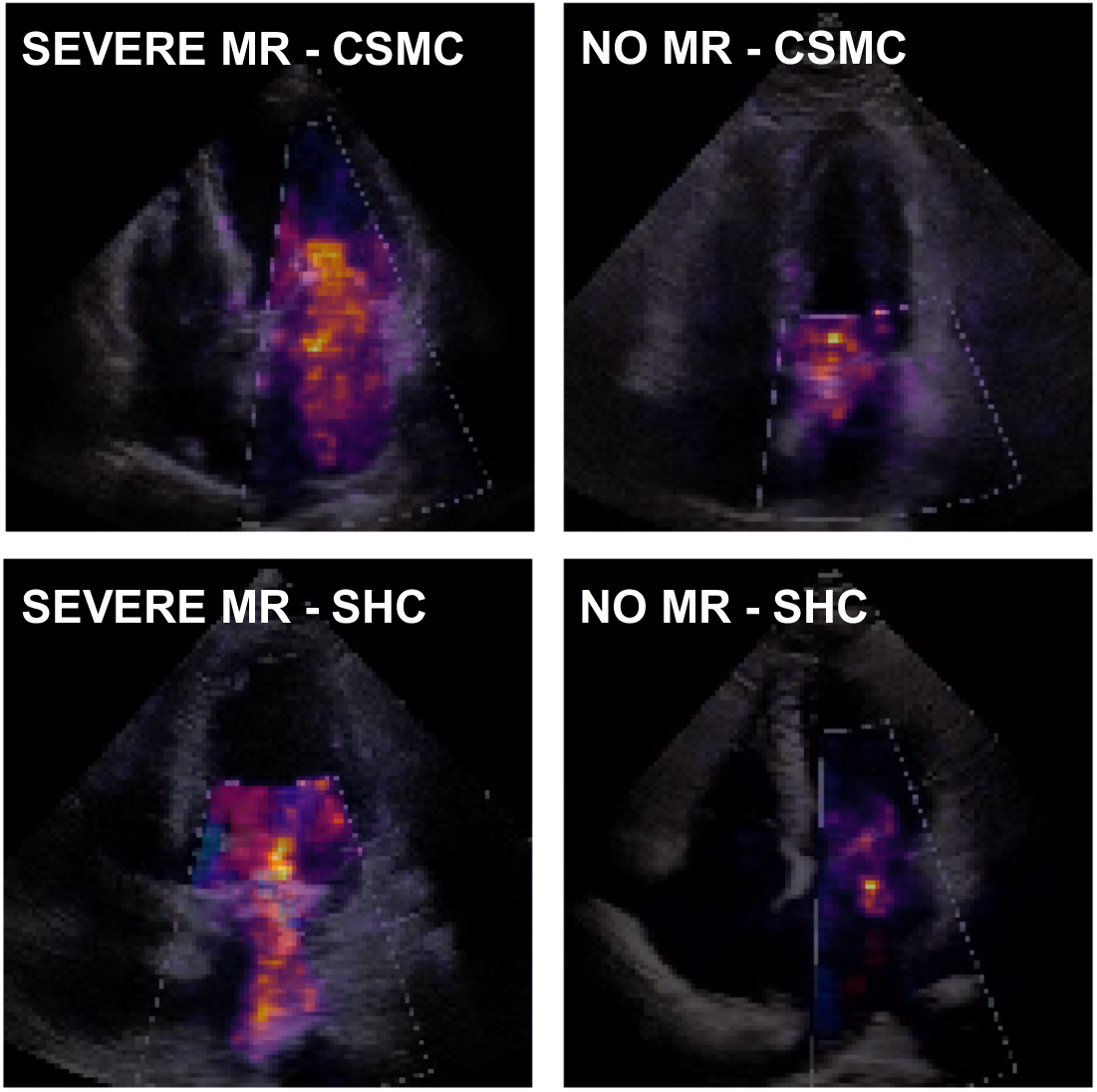
Saliency Map Visualization for MR classification models. Echocardiogram videos with severe MR from CSMC (top left) and Stanford (bottom left) are shown on the left, while videos with no MR from CSMC (top right) and Stanford (bottom right) are shown on the right. Saliency maps were computed using the Integrated Gradients method. A final 2-dimensional heatmap was generated by using the maximum value along the temporal axis for each pixel location in the video. Pixels brighter in color and closer to yellow were more salient to model predictions, while those darker in color were less important to the model’s final prediction. Severe MR was assessed by using the activation function for severe disease to generate a heatmap. When assessing controls, heatmaps were generated by stacking heatmaps for severe and moderate classes and taking the maximum between the two at each pixel location.

## Discussion

We developed and validated an automated pipeline for assessing for mitral regurgitation in echocardiography. From a full transthoracic echocardiogram study, the algorithm automatically screens for appropriate A4C videos with color Doppler on the mitral valve and then assesses MR severity. For both severe MR as well as ≥ moderate MR, the model demonstrated strong performance (> 0.916 AUC and > 0.863 NPV). This automated workflow worked in unselected external validation studies without preselection or exclusion of other concomitant comorbidities. Given these characteristics, our deep learning model could aid in the preliminary assessment of MR, facilitate review of institutional databases, or expand access for screening in low-resource settings.

Our algorithm learns features of mitral regurgitation that generalize across variability in imaging practices in two geographically distinct sites. Many prior echocardiography AI models primarily focused on black-and-white standard 2D B-mode images, while our study focuses on the AI assessment of color Doppler videos and utilized a video-based model for the incorporation of rich temporal information, both crucial for accurate MR assessment. Incorporation of Doppler information greatly expands the opportunities for AI in echo, particularly in valve disease. In expert clinical interpretation, a variety of metrics beyond just color Doppler and views are synthesized together to come up with a holistic assessment of MR severity. Intriguingly, our AI algorithm generally results in concordant interpretations with the comprehensive clinical approach while relying only on the A4C view, suggesting there is significant overlapping information as well as dependence on the A4C view in standard clinical practice.

While promising, the present work carries limitations. Echocardiographic assessment of MR depends on appropriate images being obtained, with different views potentially maximizing the visualized regurgitant jet. This algorithm would not overcome incomplete input information and insufficient images that would result underestimation of MR. Future work could focus on automatically quantifying parameters like valve leaflet thickness, effective regurgitant orifice area, regurgitant volume and fraction.

The present work builds upon prior work in the space of echocardiography and AI. Several recent works have reported strides in computer vision and echocardiography, including automated view classification^32,33^, phenotyping of left ventricular hypertrophy^15^, assessment of LV systolic function^13^, aortic stenosis risk stratification, and detection of complex congenital heart defects.^34^ Prior work in machine learning applied to MR has primarily focused on structured data and non-deep learning approaches. The combination of our algorithm with previously published works using AI to guide novices in acquiring imaging could potentially increase access to screening of MR.^20,35^

In summary, we introduce a model to screen for and stratify mitral regurgitation severity from transthoracic echocardiogram videos. To do so, we provide a workflow for isolating mitral valve color Doppler videos and automation of MR severity assessment. The models were evaluated to have good performance in internal and external test cohorts. The use of such a model, with a high AUC, NPV, and generalizability across sites, can open the door for screening of mitral valve disease in the primary care setting or in low-resource environments.

## Data Availability

The dataset of videos used in this study is not publicly available due to its potentially identifiable nature.

## Acknowledgements

A.V. is a research fellow supported by the Sarnoff Cardiovascular Research Award. S.C. acknowledges support from the Erika J Glazer Family Foundation.

## Code Statement

Our code and model weights are available at https://github.com/echonet/MR

